# ON THE UNCERTAINTY ABOUT HERD IMMUNITY LEVELS REQUIRED TO STOP COVID-19 EPIDEMICS

**DOI:** 10.1101/2020.05.31.20118695

**Authors:** Daniel Gianola

## Abstract

COVID-19 evolved into a pandemic in 2020 affecting more than 150 countries. Given the absence of a vaccine, discussion has taken place on the strategy of allowing the virus to spread in a population, to increase population “herd immunity”. Knowledge of the minimum proportion of a population required to have recovered from COVID-19 infection in order to attain “herd” immunity, *P_crit_*, is important for formulating epidemiological policy. A method for measuring uncertainty about *P_crit_* based on a widely used package, EpiEstim, is derived. The procedure is illustrated using data from twelve countries at two early times during the COVID-19 epidemic. It is shown that simple plug-in measures of confidence on estimates of *P_crit_* are misleading, but that a full characterization of statistical uncertainty can be derived from EpiEstim, which reports percentiles only. Because of the important levels of uncertainty, it is risky to design epidemiological policy based on guidance provided by a single point estimate.

## 1 Introduction

COVID-19 evolved into a pandemic in 2020 affecting more than 150 countries that vary in size, population density, economic resources and public health systems. Given the absence of a vaccine, considerable discussion (e.g., Dowdy and D’Souza, 2020) at various society levels has taken place on a strategy of allowing the virus to spread in a population, to increase population “herd immunity”. Sweden’s adoption of the approach has been at the center of the debate.

Kwok et al. (2020) estimated the minimum proportion of a population required to have recovered from COVID-19 infection in order to attain “herd” immunity, *P_crit_* in various countries. Knowing *P_crit_* is important for formulating epidemiological policies tailored to a specific country. The parameter *P_crit_* is inferred from an estimate of *R_t_*, the viral reproductive, defined as the number of infections per infector at a given time of the epidemic at time t. The formula relating these two quantities is 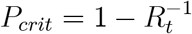 (Anderson and May, 1992). Since *P_crit_* is a fraction, it must take values between 0 and 1, implying that *R_t_* must be larger than 1 for the expression to be meaningful. As *R_t_* increases beyond 1, so does the minimum fraction of the population that needs to be infected. Since *R_t_* must be estimated soon after the epidemic starts, often there is large statistical uncertainty associated with the estimate, especially in countries where there are a few cases at the onset. Because the uncertainty about *R_t_* propagates into *P_crit_* it is important to consider it.

The objective here is to show how to measure uncertainty about *P_crit_*. The procedure is illustrated using data from twelve countries at two early times during the COVID-19 epidemic.

## 2 Inference of *P_crit_*

*R_t_* is estimated from a time series containing the number of infections *I*_0_, *I*_1_,…, *I_t_* (*t* denotes time) and from information on the statistical distribution of the virus serial interval, defined as the number of days mediating between the appearance of symptoms in pairs of infectors and infected persons. Nishiura et al. (2020) used data on 28 pairs of patients in the context of SARS-CoV-2 infections and found that a log-normal distribution with mean and standard deviation of 4.7 and 2.9 days, respectively, provided the best fit.

Table 1 of Kwok et al. (2020) displays estimates of *R_t_* and of *P_crit_* obtained at early stages of the COVID-19 epidemic for various countries. It does not report statistical uncertainty associated with the latter, although confidence intervals are provided for *R_t_*. The confidence intervals can be wide, especially for countries with smaller number of cases in the time series. For example, in the table, the width of the 95% confidence interval on *R_t_* for the USA is 0.28 whereas for Slovenia it is 3.47. A rough confidence interval for *P_crit_* can be constructed by a plug-in method using values of the bands for *R_t_*, but this way of proceeding may be misleading.

**Table 1:**
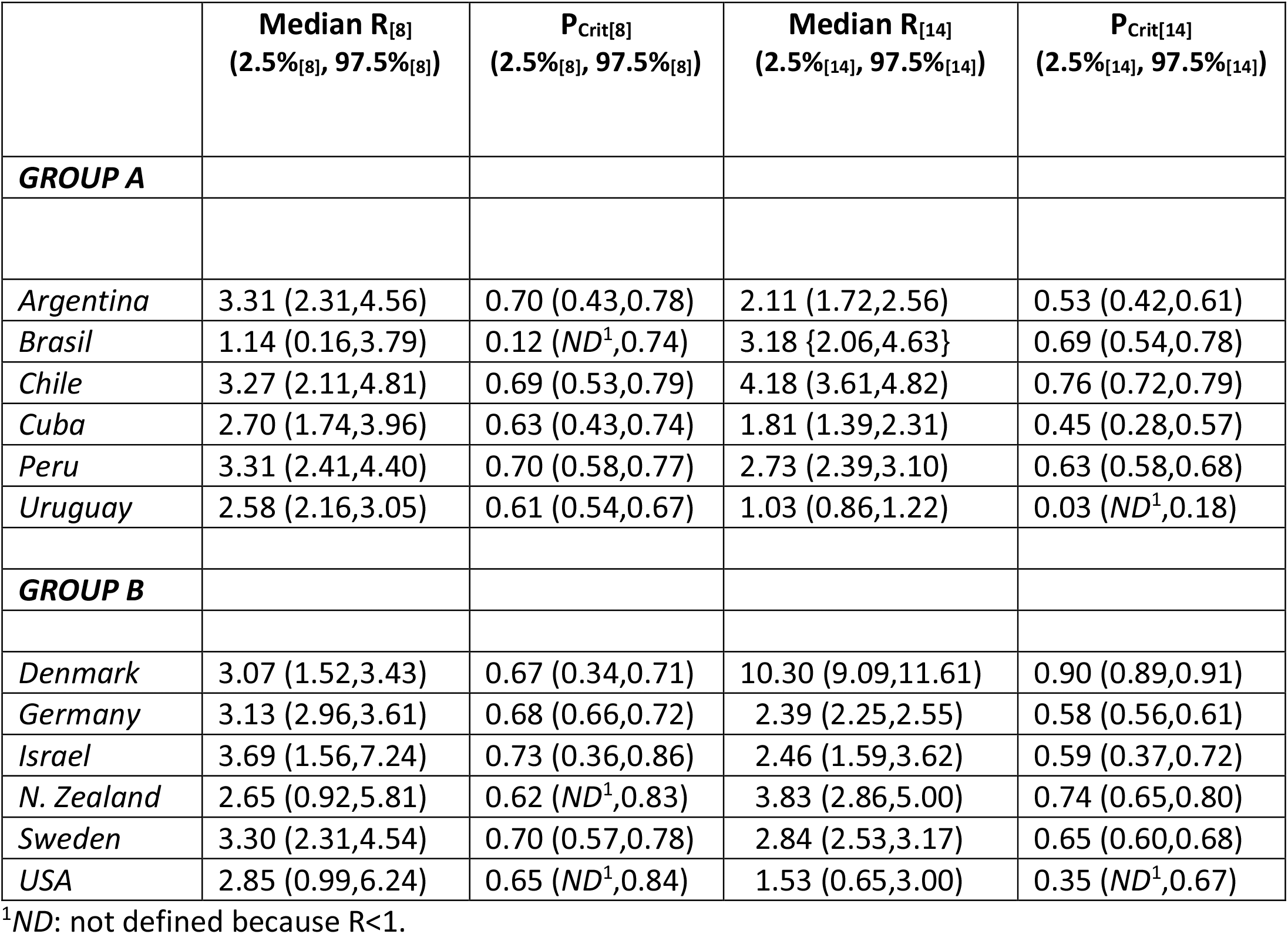
Estimates of the median, 2.5 and 97.5 percentiles of the posterior distribution of R_[t]_ (virus reproducing number) at days t=8, 14 of the COVID-19 pandemic in twelve countries. The “plug-in” P_crit_ (minimum proportion of the population that needs to be infected in order to reach herd immunity) estimates are directly obtained from the relationship P_crit_ =1-R_[t]_^−1^.

A suitable measure of the statistical uncertainty associated with inferring *P_crit_* is obtained using a Bayesian approach such as the one employed in the `R` package `EpiEstim` described in Cori et al. (2013), but with some minor external (to the program) modifications. Briefly, the package employs a model that assumes that the number of infections observed at time *t* follows a Poisson distribution with parameter *R_t_*Λ_*t*_. Here, Λ_*t*_ is a weighted average of past infections; weights are calculated from knowledge (or estimation) of the serial interval distribution. If, *R_t_* is assigned a *Gamma*(*a, b*) prior distribution, the posterior is also a *Gamma*(*a_t_, b_t_*) distribution where *a_t_* and *b_t_* depend on *a* and *b*, on the number of past infections and on the distribution of the serial interval (Cori et al., 2013).

Since *R_t_* has a *Gamma*(*a_t_, b_t_*) posterior distribution, then 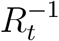 has an inverse Gamma distribution with parameters (*a_t_, b_t_*) (Sorensen and Gianola, 2002; Gelman et al., 2014) with density

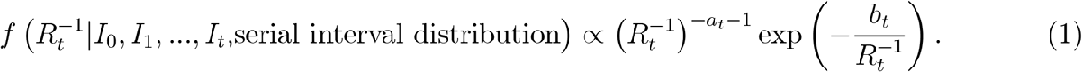

Since 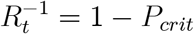 is a linear relationship, the posterior distribution of *P_crit_* has density

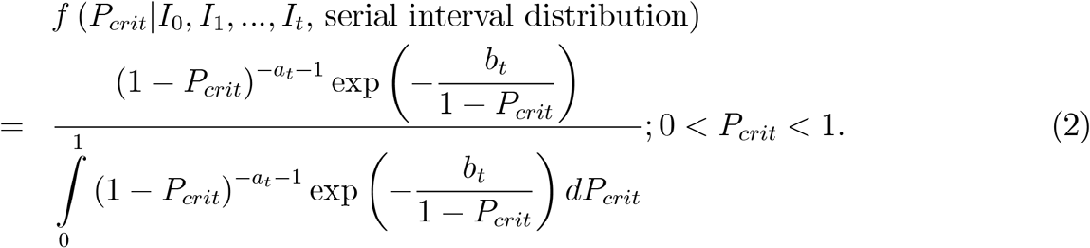

For instance, if *a_t_* = 164 and *b_t_* = 65, the posterior density of *P_crit_* is

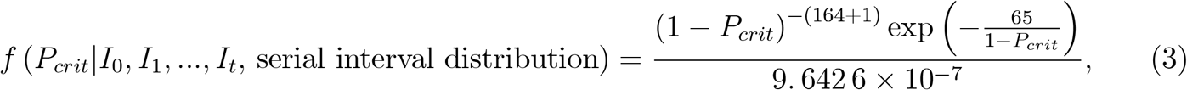

and takes the form depicted in Figure 1. The posterior expectation of 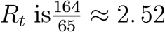 so a plug-in estimate would give 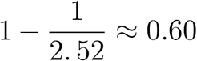 as an approximation to the posterior mean of *P_crit_*, implying that a minimum 60% of the population needs to be infected in order to attain herd immunity. The posterior mean and standard deviation of *P_crit_* are 0.60 and 0.03, respectively, and the coefficient of variation is 5.2%. The posterior probability that *P_crit_* is larger than 0.60 is 53.6%.; the probability that it is smaller than 0.55 is 5.9%, and the probability that it takes values between 0.55 and 0.60 is about 40.5%. Assuming the case fatality rate is 1%, in a country with a population of 10 million persons (e.g., Azerbaijan or Sweden), the “road” to immunity would produce about 55,000 expected deaths if *P_crit_* = 0.55, versus 62,000 if it were *P_crit_* = 0.62, a difference of 7,000 person dying. The larger the uncertainty about *P_crit_*, the larger the risk associated with choosing a strategy for control of the epidemic.

**Figure 1.**
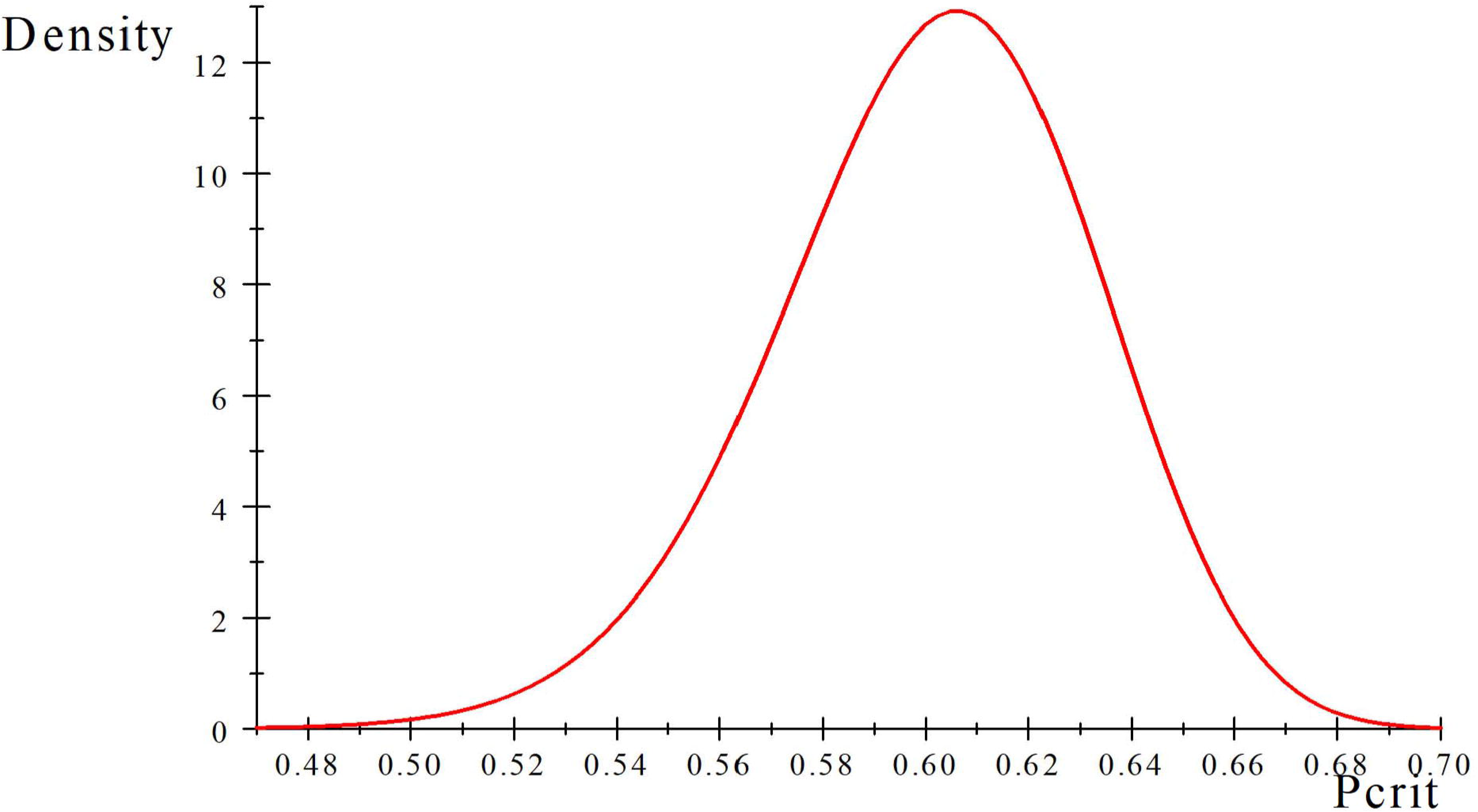
Posterior density of P(crit) for a hypothetical example. E[P(crit)] = 0.60 and the posterior coefficient of variation is 5.2%

## 3 Application to the COVID-19 pandemic

Data were downloaded from the European Centre for Disease Prevention and Control, an European Union organization based in Sweden. Twelve countries were chosen and organized into two groups as shown in Table 1. Estimates of *R_t_* (posterior median and 2.5 and 97.5 percentiles of the posterior distribution) were obtained using `EpiEstim` at times *t* = 8, 14 from the first case. Simple “plug-in” estimates of *P_crit_* and of percentiles were derived from the relationship between the latter and *R_t_*. In some cases (Brasil, Uruguay, New Zealand and USA) some percentiles could not be calculated because *R_t_* was smaller than 1, producing “plug-in” estimates of *P_crit_* outside of its parameter space. Inter-percentile ranges were wide in many instances. For example, at *t* = 8 (the earliest time at which `EpiEstim` produced an estimate of *R_t_*, the range was 35% in Argentina, 31% in Cuba, 37% in Denmark and 22% in Israel; at *t* = 14 the range was still wide for Israel, at 35%. For Sweden, the ranges were 21% and 8% at times *t* = 8 and *t* = 14, respectively. The uncertainty about *P_crit_* was large for many countries.

Instead of using the crude “plug-in”method, which can yield estimates outside of the 0 — 1 space for a proportion, `EpiEstim` can be “tricked” to produce the entire posterior distribution. At any *t*, `EpiEstim` returns the mean and standard deviation (*sd*) of the posterior distribution. If the parameterization is *Gamma*(*shape* = *a, rate* = *b*), then 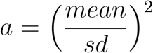 and 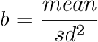 from which (2) can be numerically evaluated. Alternatively, the posterior distribution of *P_crit_* can be estimated by drawing random numbers from a computer as follows: 1) sample a large number of *Gamma*(*a, b*) deviates. 2) For each draw, form 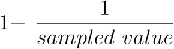, and accept it as draw from the posterior distribution of *P_crit_* if it resides in the (0, 1) interval; discard it otherwise.

The posterior densities for Group A and B countries are in Figures 2 and 3, respectively. The “plug-in” measures of uncertainty for *P_crit_* derived from the percentiles of the posterior distribution of *R_t_* were often misleading or useless. For example, for Argentina (Figure 2, curve in red) the naive lower bounds for *t* = 8 does not inform that *P_crit_* should probably be at least 50% to attain herd immunity. For Israel (Figure 3) the naive bounds exaggerate uncertainty as the posterior densities are reasonably sharp, both at *t* = 8 and *t* = 14. In Uruguay (Figure 2), the low number of cases observed translated into low values of *R_t_*, especially at *t* = 14 but suggested that herd immunity could be reached with a low proportion of the population infected, which would perhaps take a considerable amount of time but at a low cost in terms of number of dead people. However, at *t* = 8 the analysis suggested that *P_crit_* would need to be around 50 – 60%. For Sweden (Figure 3), it appears that *P_crit_* would need to be at least 55%; for the USA (Figure 3), the uncertainty was enormous, as suggested by the analyses conducted a *t* = 8 and *t* = 14.

**Figure 2.**
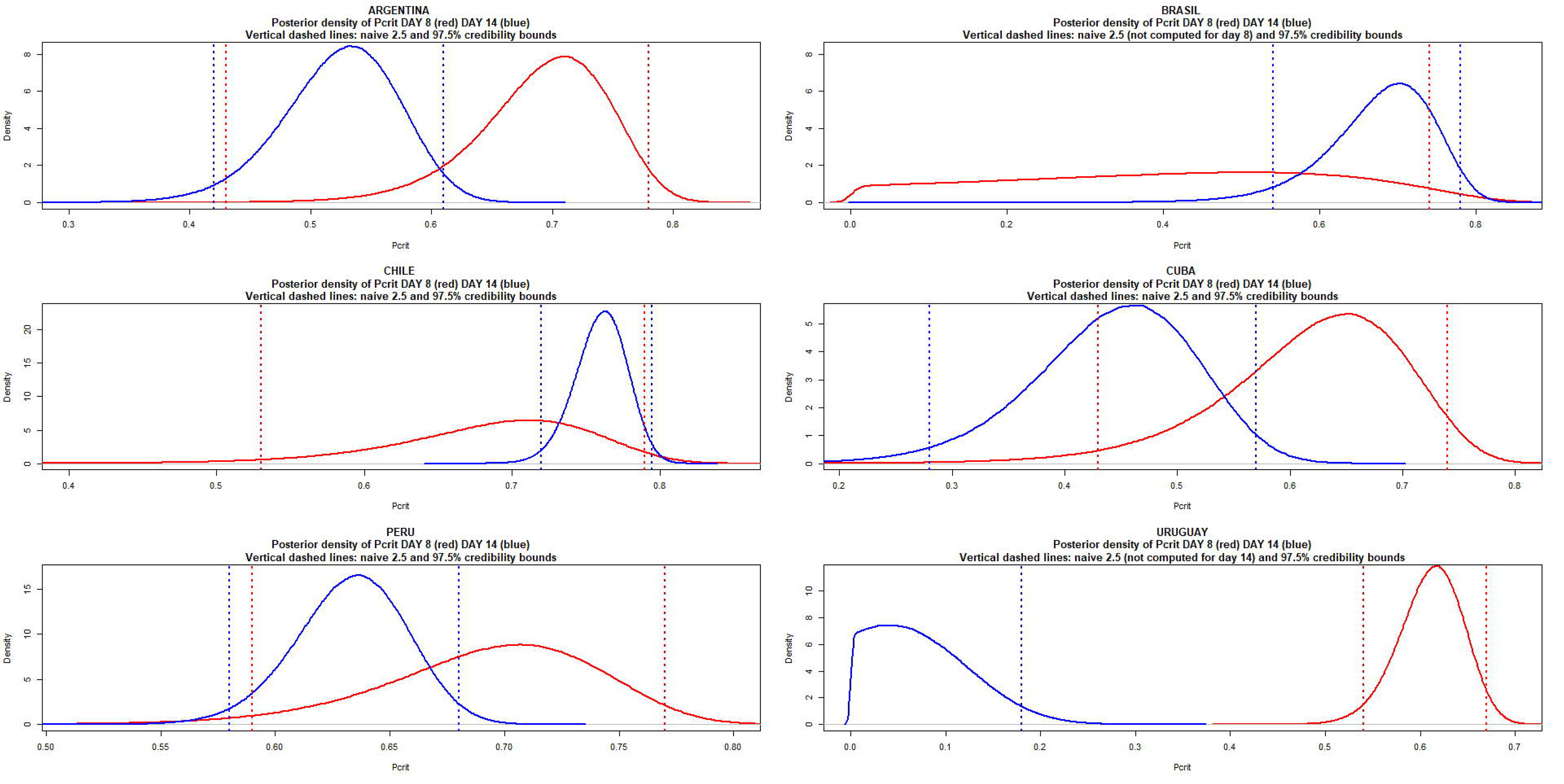
Posterior densities of P(crit) at times = 8 and 14 of the COVID19 pandemic for Argentina, Brasil, Chile, Cuba, Peru and Uruguay

**Figure 3.**
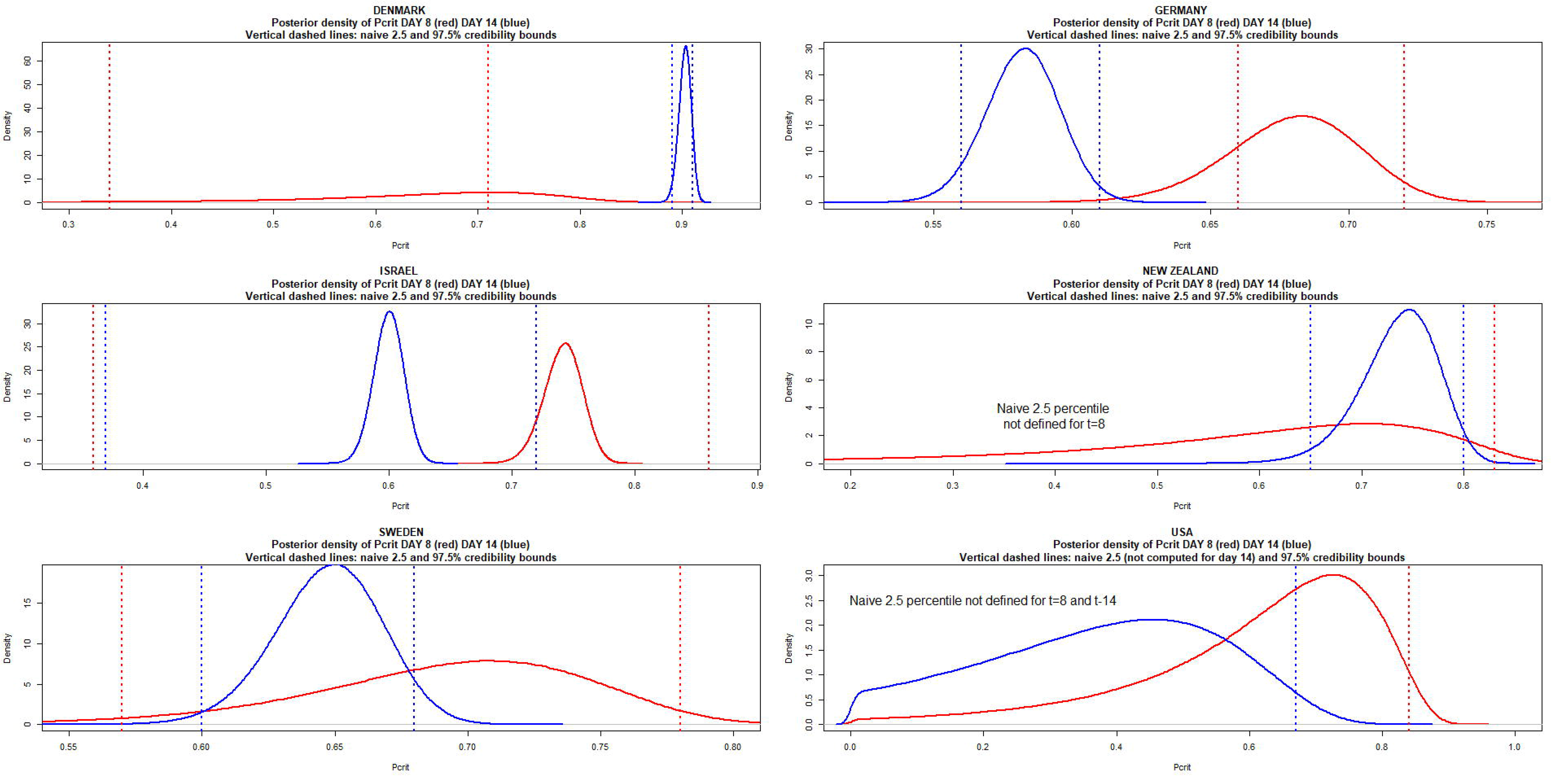
Posterior densities of Pcrit at times = 8 and 14 of the COVID19 pandemic for Denmark, Germany, Israel, New Zealand, Sweden and USA

## 4 Conclusion

Attaining herd immunity requires a minimum proportion of the population to infected. Such proportion can be inferred from estimates of the virus reproducing number calculated at early stage of the epidemic. Plug-in measures of confidence on estimates were misleading, but a full characterization of statistical uncertainty was derived from publicly available software. Because of the important levels of uncertainty, it is extremely risky to design epidemiological policy based on the guidance provided by a single point estimate, as in Kwok et al. (2020).

## Data Availability

Data are available from https://www.ecdc.europa.eu/sites/default/files/documents/COVID-19-geographic-disbtribution-worldwide

